# *IN-UTERO* MOTHER-TO-CHILD SARS-CoV-2 TRANSMISSION: viral detection and fetal immune response

**DOI:** 10.1101/2020.07.09.20149591

**Authors:** Claudio Fenizia, Mara Biasin, Irene Cetin, Patrizia Vergani, Davide Mileto, Arsenio Spinillo, Maria Rita Gismondo, Francesca Perotti, Clelia Callegari, Alessandro Mancon, Selene Cammarata, Ilaria Beretta, Manuela Nebuloni, Daria Trabattoni, Mario Clerici, Valeria Savasi

**Author notes:** corresponding author, Unit of Obstetrics and Gynecology, Department of Biomedical and Clinical Sciences, ASST Fatebenefratelli Sacco, University of Milan, via G.B. Grasi 74, 20157, Milan, Italy. The authors equally contributed to the manuscript (Claudio Fenizia, Mara Biasin).

## Abstract

Pregnancy is known to increase the risk of severe illnesses in response to viral infections. Therefore, the impact of SARS-CoV-2 infection during gestational ages might be detrimental and the potential vertical transmission should be thoroughly studied.

Herein, we investigated whether SARS-CoV-2 vertical transmission is possible and, in case, whether this results in a fetal involvement. Additionally, we analyzed the role of the antibody and the inflammatory responses in placenta and plasma from SARS-CoV-2-positive pregnant women and fetuses.

31 SARS-CoV-2 pregnant women were enrolled. Real-time PCR was performed to detect the virus on maternal and newborns’ nasopharyngeal swabs, vaginal swabs, maternal and umbilical cord plasma, placenta and umbilical cord biopsies, amniotic fluids and milk. Maternal and umbilical cord plasma, and milk were tested for specific anti-SARS-CoV-2 antibodies. RNA expression quantification of genes involved in the inflammatory response was performed on four selected placentas. On maternal and umbilical cord plasma of the same subjects, secreted cytokines/chemokines were quantified.

SARS-CoV-2 is found in at-term placentae and in the umbilical cord blood, in the vaginal mucosa of pregnant women and in milk. Furthermore, we report the presence of specific anti-SARS-CoV-2 IgM and IgG antibodies in the umbilical cord blood of pregnant women, as well as in milk specimens. Finally, a specific inflammatory response is triggered by SARS-CoV-2 infection in pregnant women at both systemic and placental level, and in umbilical cord blood plasma.

Our data strongly support the hypothesis that in-utero vertical transmission is possible in SARS-CoV-2 positive pregnant women. This is essential for defining proper obstetric management of COVID-19 pregnant women, or putative indications for mode and timing of delivery.

## INTRODUCTION

COVID-19 pandemic is currently spreading worldwide. The number of confirmed cases currently exceeding 11.5 million people, about 590,000 deaths, and Italy representing one of the most affected countries^1–3^. Severe COVID-19 cases exhibit a dysfunctional immune response characterized by higher blood plasma levels of IL-1β, IL-2, IL-6, IL-7, IL-10, granulocyte colony-stimulating factor (G-CSF), IP-10 (CXCL10), MCP1 (CCL2), MIP1α (CCL3) and tumor necrosis factor (TNF), that mediate widespread lung inflammation and fail to successfully eradicate the pathogen^1,4–7^.

Maternal physiological adaptations to pregnancy are known to increase the risk of developing severe illness in response to viral infections, such as influenza; preliminary data suggest that the prognosis of SARS-CoV-2 infection could be more severe as well in pregnant women^8^. Vertical transmission of SARS-CoV and MERS, the two other animal coronaviruses known to infect humans, was never documented to occur. However, the number of reported cases of infected pregnant women was very low and not sufficient to draw firm conclusions (12 reported cases for SARS-CoV and 11 for MERS)^9,10^. Conversely, as the number of SARS-CoV-2-positive patients is rising worldwide, multiple reports focus on SARS-CoV-2-positive pregnant women^11–16^. No trace of the virus was detected by real-time PCR, so far^s11,12,14,15,17^; however, two independent manuscripts described elevated SARS-CoV-2-specific IgG and IgM antibody levels in the blood of three newborns of SARS-CoV-2 infected mothers^18,19^. As IgG, but not IgM, are normally transferred across the placenta, this is suggestive of *in-utero* infection^18,19^. Moreover, placental sub-membrane and cotyledon was reported positive to the virus in a 20 weeks miscarriage of a SARS-CoV-2-positive pregnant woman^20^. As recently reported, the two known SARS-CoV-2 receptors Angiotensin Converting Enzyme 2 (ACE2) and Transmembrane Protease Serine 2 (TMPRSS2) are widely spread in specific cell types of the maternal-fetal interface^21^. Therefore, the impact of the virus on placenta and the potential of vertical transmission of SARS-CoV-2 need to be further carefully addressed.

Herein we investigated: 1) whether SARS-CoV-2 vertical transmission is possible, 2) how the production of antibodies occurs in the mother and possibly in the fetus and 3) the inflammatory profile in COVID-19-positive pregnant women and fetuses. Answering these questions is essential for understanding virus biological behavior during pregnancy, and for defining proper obstetric management of COVID-19 pregnant women.

## MATERIALS AND METHODS

### Study population

This is a prospective multicenter study that includes 31 women: 30 with laboratory-confirmed COVID-19 infection admitted at delivery in three COVID-19 maternity hospitals of Lombardy, Italy: the ‘L. Sacco’ Hospital (Milan), the S. Gerardo Hospital/MBBM Foundation (Monza), and the San Matteo Hospital (Pavia) between March 9 and April 14, 2020. A further woman, (subject n. 31), the wife of Italian patient One, was found to be SARS-CoV-2-positive at the 32 gestational weeks and delivered at Buzzi maternity Hospital, a COVID-19-free hub and was admitted in the study as well.

All women underwent clinical evaluation of vital signs and symptoms, laboratory analysis and radiological chest assessment at admission at discretion of physicians. The therapeutic management was consequently tailored according to the clinical findings and national guidelines^22^. Demographic and anthropometric characteristics, medical and obstetric comorbidities, were recorded at enrollment through a customized data collection form. All pregnancies were singleton, with a normal course and regular checks, until delivery.

Data on mode of delivery or pregnancy termination, maternal and neonatal outcomes, and postpartum clinical evolution (e.g. breastfeeding, skin to skin, etc.) were subsequently recorded. Data accuracy was independently verified by two study investigators.

The protocol was approved by the local Medical Ethical and Institutional Review Board.

### Specimen collection

Biological samples were collected at admission (T0), delivery (T1) and post-partum (T2), as summarized in figure 1. T0 samples included a nasopharyngeal swab in order to test the positivity for SARS-CoV-2. At T1, full thickness placental and umbilical cord biopsies were obtained and a 10 ml umbilical cord blood sample was collected in EDTA after cleaning throughout the cord with a sterile gauze and physiological solution before sampling. Both biopsies and blood samples were obtained in sterile way by a dedicated operator. In case of caesarean section, if possible, amniotic fluid was collected. Moreover, a 10 ml maternal blood sample in EDTA was collected, together with a vaginal swab before labor or cesarean section. Upon delivery, a nasopharyngeal swab was immediately performed on newborns and mothers. Approximately five days after delivery (T2), transitional breastmilk samples were collected from all breastfeeding women. For each subject, days occurring between T0 and T1 (ΔT1-T0) were calculated. Fourteen sets of samples from the Obstetrics and Gynecology Unit of “L. Sacco” Hospital (Milan) were immediately transferred to the dedicated laboratory of Clinical Microbiology, Virology and Diagnostics, “L. Sacco” Hospital, and/or to the laboratory of Immunology, University of Milan, according to the kind of specimen, to be readily processed. Seventeen samples collected at S. Gerardo Hospital/MBBM Foundation (Monza), and San Matteo (Pavia) were frozen at −80°C upon collection and transferred to the same laboratories in dry ice.

**Figure 1.**
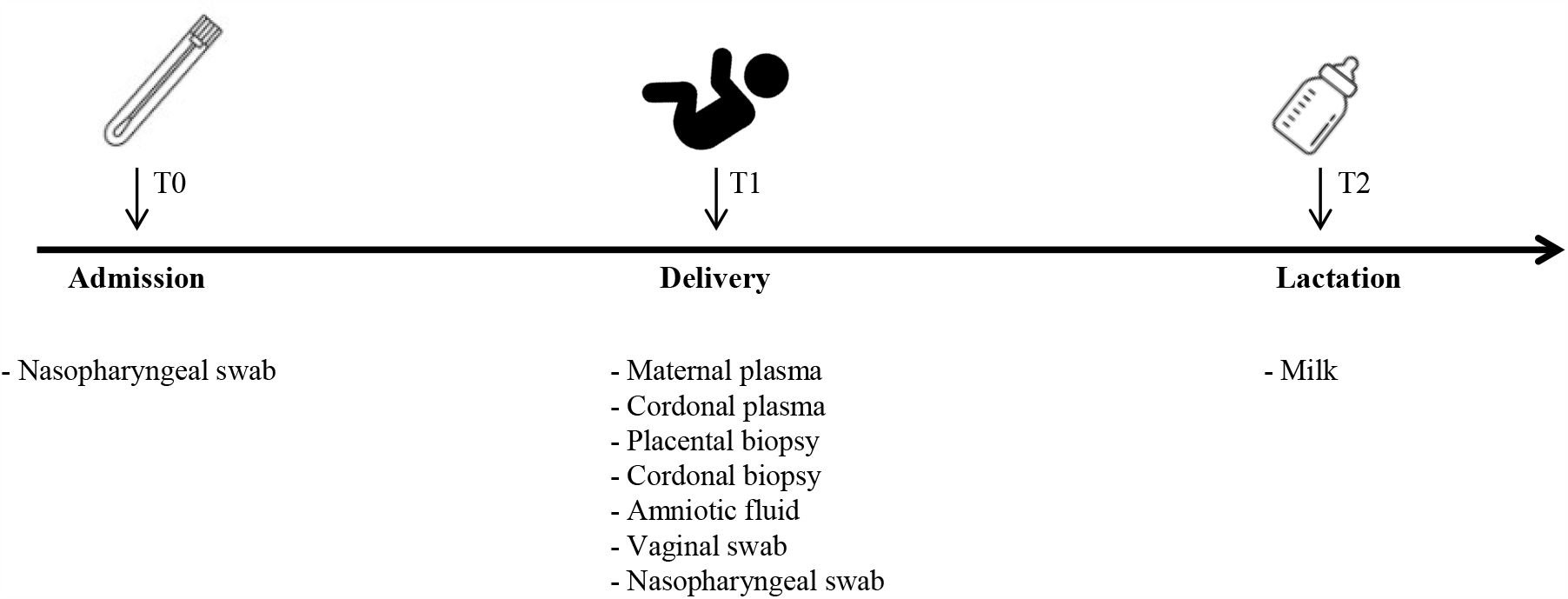
Specimen collection timeline. List of specimens collected at admission to the study (T0), at delivery (T1) and during lactation (T2) for each enrolled subject.

### Diagnostic analyses

Molecular analysis was performed to detect viral RNA, using the automated Real-Time PCR ELITe InGenius® system and the GeneFinderTM COVID-19 Plus RealAmp Kit assay (ELITechGroup, France). The reaction mix was prepared according to manufacturer’s instruction. Three target genes, RNA-dependent RNA polymerase (RdRP), Nucleocapsid (N) and Envelope (E) have been simultaneously amplified and tested. A cycle threshold value (Ct-value) lower than 40 was defined as a positive test result according to the manufacturer’s instruction.

The presence of SARS-CoV-2 specific antibodies was investigated using SARS-CoV-2 IgG and IgM chemiluminescence immunoassay (CLIA) kits on fully automated iFlash1800 analyzer (Shenzen YHLO Biotech Co., Ltd., Shenzen, China): the assay uses Nucleocapsid (N) and Spike (S) viral proteins as magnetic beads coating antigens. The value of 10.0 AU/mL was used as positivity cut-off for IgM, while 7.1 for IgG^23^.

### Tissues processing

Placental and umbilical cord biopsies were manually dissected into few sections of approximately 2mm^3^. Such sections were then thoroughly homogenized and total RNA was isolated using the acid guanidium thiocyanate–phenol– chloroform method (RNAbee, Duotech, Milan, Italy), as previously described^24^. Alternatively, biopsies were paraffin-embedded and stored as such.

Plasma samples were collected from blood of all the enrolled subjects, as well as plasma samples from funicular blood, amniotic fluid and vaginal swabs. Moreover, as control for molecular analyses, plasma from a SARS-CoV-2 negative pregnant woman (CTR-), as well as plasma samples from funicular blood and placental tissues were included. RNA was extracted by the Maxwell® RSC Instrument with Maxwell® RSC Viral Total Nucleic Acid Purification Kit (Promega, Fitchburg, WI, USA). As result, RNA eluted in RNAse-free water was obtained.

Once RNA was reverse transcribed into cDNA^24^, real-time PCR was performed on a CFX96 (Bio-rad, CA, USA) using TaqMan probes specifically designed to target two regions of the nucleocapsid (N) gene of SARS-CoV-2. For such application, we employed the 2019-nCoV CDC qPCR Probe Assay emergency kit (IDT, Iowa, USA) which include also primers and probes that target the human RNase P gene.

### Expression analyses of inflammatory response

The inflammatory response was analysed in four selected RNA samples extracted from placenta biopsies of one negative control (CTR-), one SARS-CoV-2 recovered (subject n. 31) subject and two SARS-CoV-2 positive subjects (subjects n. 17 and 25). Subjects n. 17 and 25 gave birth to SARS-CoV-2 positive newborns, according to the first nasopharyngeal swab (T1). Analyses were performed by a PCR array that include a set of 84 optimized real-time PCR primers plus 5 housekeeping genes on a 96-well plates; the procedures suggested by the manufacturer were followed (Qiagen, Hilden, Germania). Undetermined raw CT values were set to 35. Only variables with at least a two-fold increase in their value are presented and discussed in the manuscript.

Concentration of 27 cytokines was assessed in maternal and funicular plasma samples from the very same four subjects using immunoassays formatted on magnetic beads (Bio-rad, CA, USA) according to manufacturer’s protocol via Luminex 100 technology (Luminex, Texas, USA).

### Statistics

For the study variables, medians and ranges were reported for quantitative variables. The analyses were performed using SPSS Statistics, Version 26.0 (IBM Corp. Armonk, NY) together with GraphPad Prism 8.

All the procedures were carried out in accordance with the GLP guidelines adopted in our laboratories.

## RESULTS

### Population

Four patients were classified as severe cases, defined by the need of urgent delivery for the deterioration of maternal conditions or by ICU/sub-intensive care admission. A radiological confirmation of interstitial pneumonia was obtained on admission or antepartum for all the severe cases and in 10 (32%) of the mild cases. Pharmacological treatment during the antepartum period of hospitalization is reported in table 1. In the only severe case of pre-term labor (subject n. 17), corticosteroids for RDS-prophylaxis were administered.

**Table 1.** Baseline characteristics of the study population on admission.

| **CHARACTERISTICS AT DELIVERY** | **Total study population**<br>**(n=31)** |
| --- | --- |
| **MATERNAL BASELINE CHARACTERISTICS** |  |
| **Maternal age**, years, median (range) | 30 (15-45) |
| **RT-PCR assay of a maternal nasopharyngeal swab** |  |
| Positive n (%) | 30 (97) |
| Negative n (%) | 1* (3) |
| **Prepregnancy BMI**, kg/m<sup>2</sup>, median (range) | 23 (17-37) |
| **Known sick contact**, n (%) | 10 (32) |
| **Smoking habit**, n (%) | 0 |
| **Ethnicity**, Caucasian, n (%) | 21 (68) |
| **Chronic comorbidity**, n (%) | 11 (35) |
| **Parity**, nulliparous n (%) | 14 (45) |
| **Flu vaccination in pregnancy**, n (%) | 8 (26) |
| **ANTE-PARTUM THERAPY** |  |
| Antibiotic, n (%) | 10 (32) |
| antiviral, n (%) | 8 (26) |
| hydroxychloroquine, n (%) | 13 (42) |
| oxygen support without ICU admission, n (%) | 4 (13) |
| **POSITIVE CHEST X-RAY**, n (%) | 14 (45) |
| **SEVERE CASE**, n (%) | 4 (13) |
| **ADMISSION TO ICU**, n (%) | 1 (3) |

\* subject n. 31, SARS-CoV-2 recovered at the time of enrolment.

Maternal and pregnancy outcomes in the study population are reported in Table 2. Regarding mode of delivery, three patients underwent emergency deliver for maternal respiratory indication. Among the severe cases, one needed post-partum admission to ICU and invasive ventilation for eleven days in total.

**Table 2.** Maternal and pregnancy outcomes in the study population.

|  | **Total study population**<br>n=31 |
| --- | --- |
| **Delivery mode** |  |
| - vaginal, n (%) | 25 (81) |
| - caesarean section, n (%) | 6 (19) |
| **GA at delivery**, weeks median (range) | 39 (34.4-41.4) |
| **Induction of delivery related to COVID-19**, n (%) | 6 (19) |
| **Caesarean section for severe maternal illness related** | 3 (9) |
| **COVID-19**, n (%) |  |
| **Preterm delivery**, n (%) | 1 (3) |
| **Fetal gender**, male, n (%) | 18 (58) |
| **Birth weight**, g, median (range) | 3200 (2180- 4165) |
| **Umbilical artery pH**, median (range) | 7.31 (7.14-7.43) |
| **APGAR score 5'** < 7, n (%) | 1 (3) |
| **Newborn COVID**, positive, n (%) | 2 (6) |
| **NICU admission**, n (%) | 2 (6) |
| **Skin to Skin**, n (%) | 4 (13) |
| **Breastfeeding**, n (%) | 29 (94) |

Subject n. 31 became negative at week 35 of pregnancy and delivered spontaneously at week 38. Except in one case (subject n. 17), all pregnancies were full term. Subject n. 17 was admitted preterm at 33+6 weeks with fever and dyspnea and delivered spontaneously at 34+4 weeks. A female baby was born, weighing 2180 g, with an Apgar Score of 9 and 10 at 1 and 5 minutes, respectively, with a pH of umbilical artery of 7.14. The newborn was diagnosed with COVID-19 infection through a nasopharyngeal swab and was admitted to NICU for prematurity. Subject n. 25 spontaneously delivered at week 39+2. A male baby was born, weighing 3340 g, with an Apgar Score of 9 and 10 at 1 and 5 minutes, respectively, and the umbilical artery pH of was 7.14. The newborn was diagnosed with SARS-CoV-2 infection through a nasopharyngeal swab upon delivery, while he tested negative 48h later. Except for the two above-mentioned cases, no other newborns resulted positive to SARS-CoV-2 detection by nasopharyngeal swab. Except for two cases, all newborns were breastfed. All the newborns were healthy, including the two SARS-CoV-2 positive ones.

### Virus and antibody detection

We investigated the presence of SARS-CoV-2 in the collected specimens, as shown in Table 3 and summarized in Table 4a. We detected the virus in two (6%) maternal plasma samples (subjects n. 4 and 17), both of them characterized by a severe clinical outcome. Remarkably, we detected the presence of SARS-CoV-2 in vaginal swab, placental tissue and cord plasma from subject n. 17. Moreover, we detected SARS-CoV-2 in placental tissue from subject n. 25. We detected SARS-CoV-2 in one milk specimen only, from a severe clinical outcome case (subject n. 1). None of the tested six amniotic fluids, nor the twelve umbilical cords, resulted positive (Table 3 and 4a).

**Table 3.** Synoptic table of maternal and fetal SARS-CoV-2 virus and anti-SARS-CoV-2 antibody detection correlated to clinical data.

| subject n. | clinical outcome | $\Delta$ T1-T0 (days) | maternal plasma | | | vaginal swab | Placenta | Umbilical cord plasma | | | umbilical cord | milk | | |
| --- | --- | --- | --- | --- | --- | --- | --- | --- | --- | --- | --- | --- | --- | --- |
|  |  |  | virus | IgM | IgG |  |  | virus | IgM | IgG |  | virus | IgM | IgG |
| 1 | SEVERE | 2 | - | - | - | - | - | - | - | - | - | + | + | - |
| 2 | MILD | 1 | - | - | - | - | - | N/A | N/A | N/A | - | - | - | - |
| 3 | MILD | 1 | - | - | - | - | - | - | - | - | - | - | - | N/A |
| 4 | SEVERE | 2 | + | - | + | - | - | - | - | - | - | - | - | - |
| 5 | MILD | 7 | - | - | - | - | - | - | - | - | - | N/A | N/A | N/A |
| 6 | MILD | 1 | - | + | + | - | - | - | - | - | - | - | N/A | - |
| 7 | MILD | 12 | - | + | + | - | - | - | - | - | - | - | - | - |
| 8 | SEVERE | 6 | - | + | + | - | - | - | - | - | - | N/A | N/A | N/A |
| 9 | MILD | 1 | N/A | N/A | N/A | - | - | - | - | - | - | - | - | - |
| 10 | MILD | 1 | - | - | - | - | - | - | - | - | - | - | - | - |
| 11 | MILD | 5 | - | - | - | - | - | - | - | - | - | - | - | - |
| 12 | MILD | 4 | - | - | + | - | - | - | - | - | N/A | N/A | N/A | N/A |
| 13 | MILD | 3 | - | - | + | - | - | - | - | + | - | - | - | - |
| 14 | MILD | 3 | - | - | + | - | - | - | - | + | N/A | N/A | N/A | N/A |
| 15 | MILD | 4 | - | - | - | - | - | - | - | - | N/A | N/A | N/A | N/A |
| 16 | MILD | 2 | - | - | - | - | - | - | - | - | N/A | N/A | N/A | N/A |
| 17 | SEVERE | 6 | + | + | + | + | + | + | - | + | N/A | N/A | N/A | N/A |
| 18 | MILD | 2 | - | - | + | - | - | - | - | + | N/A | N/A | N/A | N/A |
| 19 | MILD | 9 | - | + | + | - | - | - | - | + | N/A | N/A | N/A | N/A |
| 20 | MILD | 3 | - | + | + | - | - | - | - | + | N/A | N/A | N/A | N/A |
| 21 | MILD | 13 | - | - | + | - | - | - | - | - | N/A | N/A | N/A | N/A |
| 22 | MILD | 10 | - | - | - | - | - | - | - | - | N/A | N/A | N/A | N/A |
| 23 | MILD | 9 | - | - | - | - | - | - | - | - | N/A | N/A | N/A | N/A |
| 24 | MILD | 12 | - | - | - | - | - | - | - | - | N/A | N/A | N/A | N/A |
| 25 | MILD | 17 | - | + | + | - | + | - | + | + | N/A | N/A | N/A | N/A |
| 26 | MILD | 13 | - | + | + | - | - | - | - | + | N/A | N/A | N/A | N/A |
| 27 | MILD | 1 | - | - | + | - | - | - | - | + | N/A | N/A | N/A | N/A |
| 28 | MILD | 3 | - | + | + | - | - | - | - | + | N/A | N/A | N/A | N/A |
| 29 | MILD | 2 | - | - | + | - | - | - | - | - | N/A | - | - | - |
| 30 | MILD | 1 | - | - | + | - | - | - | - | + | N/A | N/A | N/A | N/A |
| 31 | recovered | N/A | - | - | + | N/A | - | - | - | + | N/A | N/A | N/A | N/A |

N/A = not available.
ΔT1-T0 (days) refers to the time spanning between the first COVID19 diagnosis (T0) and the delivery (T1).

**Table 4.** Summary of maternal and fetal SARS-CoV-2 virus (a) and anti-SARS-CoV-2 antibody (b) detection.

|  | **Maternal**<br>**plasma (n=30)** | **Vaginal swab**<br>**(n=31)** | **Placenta**<br>**(n=31)** | **Umbilical Cord**<br>**plasma**<br>**(n=30)** | **Umbilical cord**<br>**(n=12)** | **Amniotic**<br>**fluid**<br>**(n=6)** | **Milk**<br>**(n=11)** |
| --- | --- | --- | --- | --- | --- | --- | --- |
| **Pos, % (n)** | 6.7 (2) | 3.2 (1) | 6.4 (2) | 3.3 (1) | 0 (0) | 0 (0) | 9.1 (1) |

**b.**
|  | **Maternal plasma**<br>**(n=30)** | **Umbilical cord plasma**<br>**(n=30)** | **Milk**<br>**(n=10)** |
| --- | --- | --- | --- |
| **IgM, % (n)** | 32.1 (9) | 3.3 (1) | 10 (1) |
| **IgG, % (n)** | 63.3 (19) | 40.0 (12) | 0 (0) |
| **IgM/IgG, % (n)** | 32.1 (9) | 3.3 (1) | 0 (0) |

SARS-CoV-2-specific IgM were detected in 32% of the maternal plasma, while virus-specific IgG were present in 63% of cases. Interestingly, we detected the presence of IgM in the cord plasma in one newborn only (n. 25), whose placenta tested positive to SARS-CoV-2, while IgG were present in 40% of the umbilical cord plasma. Subject n. 1 displayed IgM in milk sample (Table 3, summarized in Table 4b).

### Increased inflammatory response in SARS-CoV-2-positive subjects

To determine whether SARS-CoV-2 infection results into an alteration of inflammatory gene expression in placenta tissue, we analysed the expression of 84 genes involved in the inflammatory response in four selected placenta biopsies. Results showed that placentae from SARS-CoV-2 infected patients (subjects n. 17 and 25) display a generalized immune activation profile compared to the uninfected one (CTR-) (Figure 2). Likewise, subject 31, who was infected at gestational week 32, but fully recovered 4 weeks before delivery, showed an increased inflammatory profile when compared to CTR-. Notably, such hyper-activation status was far more evident in placenta biopsy from subject n. 17 and even more in n. 25, whose nasopharyngeal swab tested positive to SARS-CoV-2 detection at T1, compared to the placenta biopsy from patient 31, who tested negative at the time of delivery. The genes whose mRNA expression was clearly upregulated in subjects 17 and 25 are involved in different aspects of inflammatory response and include effector cytokines and chemokines, adaptive immunity mediators, downstream signalling molecules, Toll-Like receptors (Figure 2).

**Figure 2.**
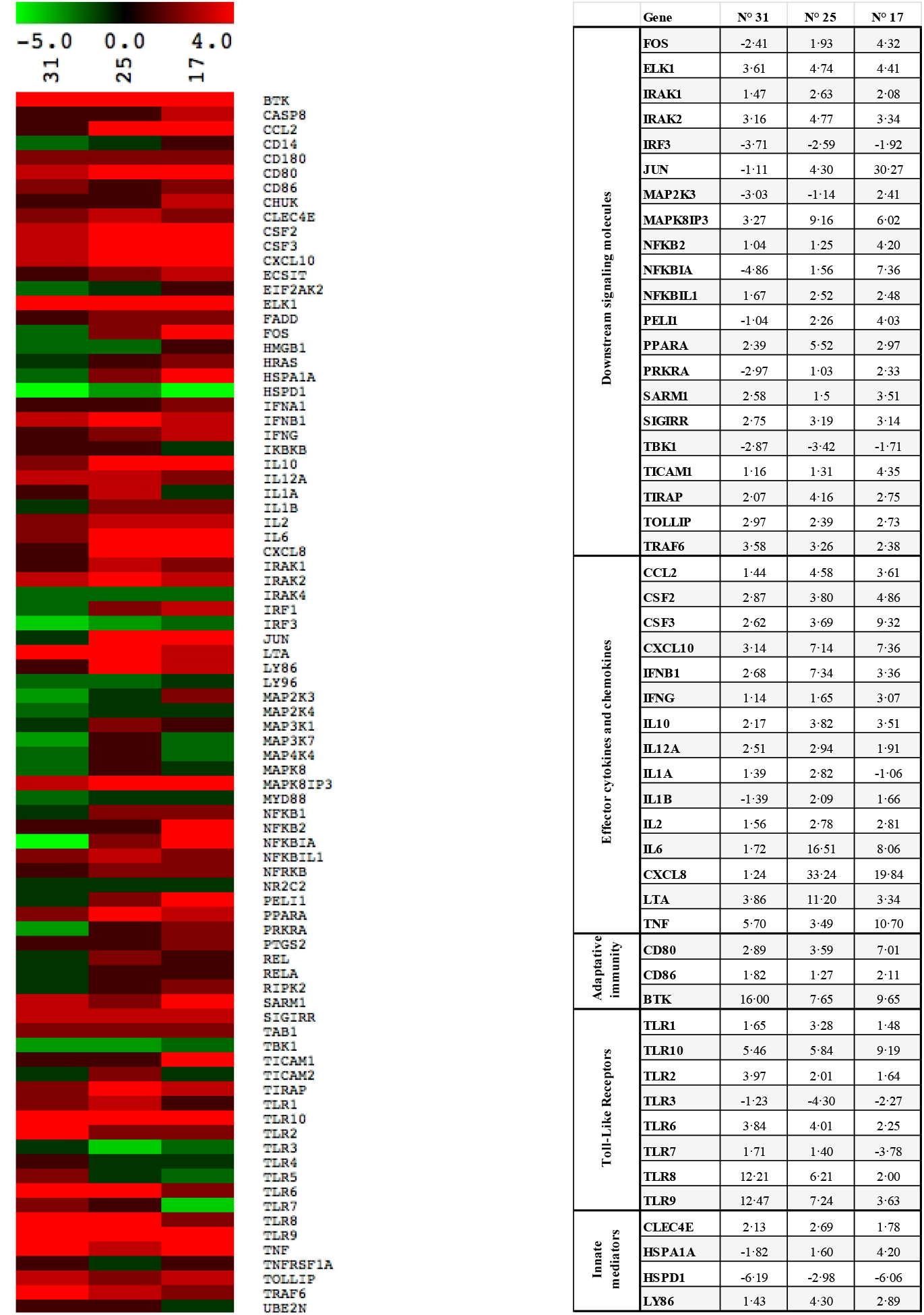
mRNA expression of 84 genes that are part of the inflammatory response is altered in placentae from subjects who experienced SARS-CoV-2 in-utero viral transmission. Real-time PCR array was performed on placenta biopsies from subjects n. 31, 25 and 17. Results are shown as a ratio of each SARS-CoV-2-positive subject compared to a SARS-CoV-2-negative one (CTR-). Gene expression (*n*fold) is shown as a color scale from green to red (MEV multiple experiment viewer software). Only targets showing at least >2-fold modulation are shown in the table.

An in-depth investigation of the cytokine/chemokine profile was carried out next in subjects CTR-, 17, 25 and 31, due to their peculiarities. A 27-cytokine multiplex assay was performed on plasma isolated from their maternal and funicular blood samples. Overall, the results obtained on maternal plasma confirmed what observed at the mRNA level in the placentae. Briefly, pro-inflammatory antiviral cytokines and chemokines were upregulated in patients n. 17 and 25, compared to subject CTR- and 31. Subject 25 displayed a more pronounced pro-inflammatory profile. In particular, IL-1β and IL-6 production were higher in subjects 25 compared to all the other subjects (Figure 3a). The same analysis was performed on funicular plasma. As for maternal plasma, the concentration of pro-inflammatory molecules was strongly increased in the newborns from subjects n. 17 and 25. Actually, this rise was mostly evident for the chemokines: IL-8, CCL2, CCL3, CCL5 and CXCL10 (Figure 3b).

**Figure 2.**
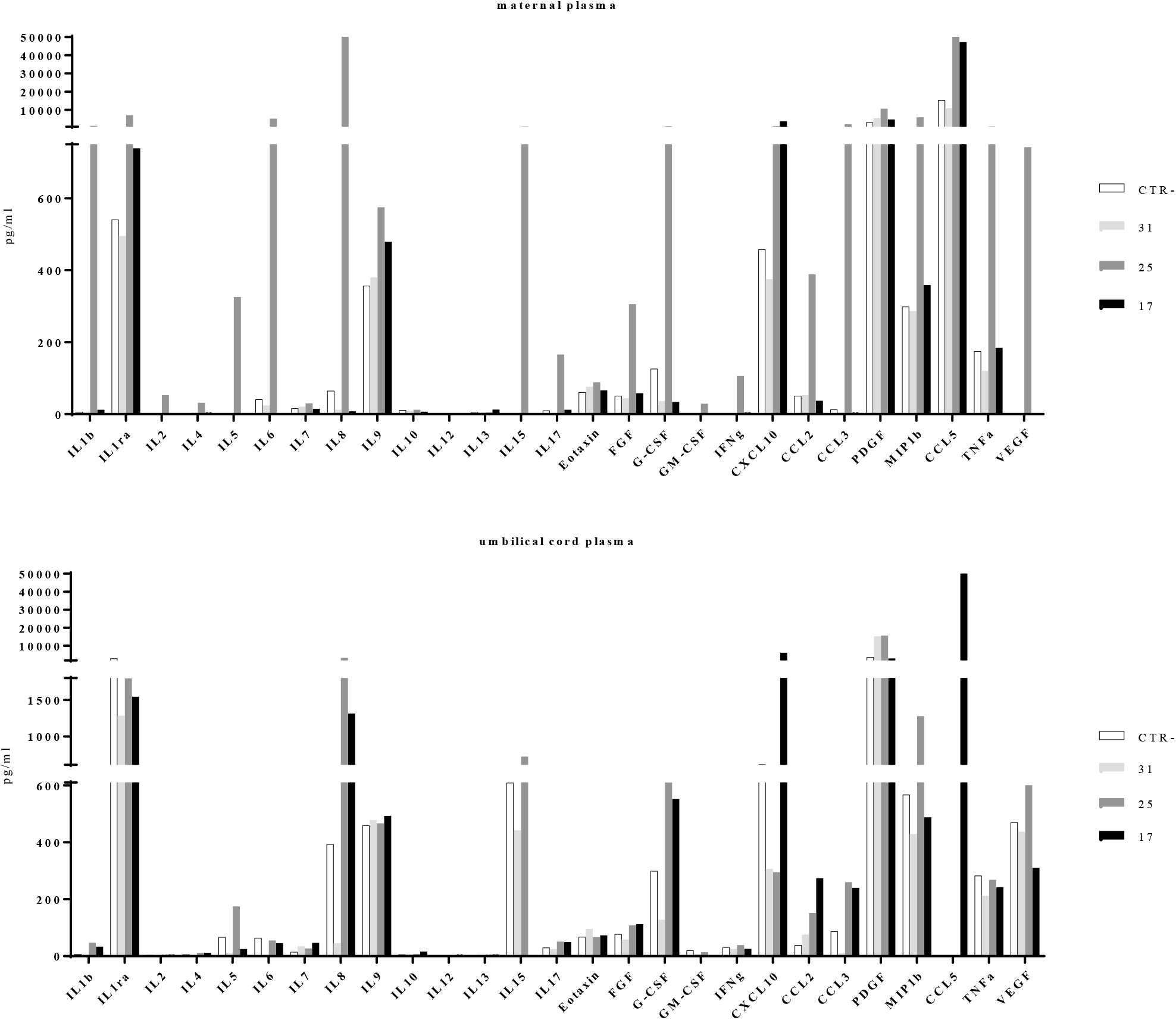
Protein secretion of 27 cytokines/chemokines that are part of the inflammatory response is altered in maternal and umbilical cord plasma from subjects who experienced SARS-CoV-2 in-utero viral transmission. Multiplex array was performed on maternal and umbilical plasma from subjects n. 31, 25 and 17. As reference, a SARS-CoV-2-negative plasma is shown (CTR-). Protein concentration is shown as pg/ml of plasma.

## DISCUSSION

We report for the first time that SARS-CoV-2 is found in the vagina of a pregnant woman, in at-term placentae and in the umbilical cord blood. Furthermore, we report the presence of specific anti-SARS-CoV-2 IgM and IgG antibodies in the umbilical cord blood of pregnant women, as well as in milk specimens. Notably, we also provide evidences that SARS-CoV-2 can be found in milk specimen. Our data support the hypothesis that *in-utero* vertical transmission is possible in SARS-CoV-2 positive pregnant women. Finally, this is the first report describing the inflammatory response triggered by SARS-CoV-2 infection in pregnant women at both systemic and placental level.

Our results strongly suggest *in-utero* vertical transmission in 2 of the 31 (6%) enrolled SARS-CoV-2 positive women. As one case was characterized by a severe clinical outcome (subject n.17), while the other one was classified as mild (subject n. 25), we speculate that the risk of mother-to-child viral transmission does not directly depend on the severity of disease progression. Supporting this observation, the clinical history as well as the results of the viro-immunological test performed on these two subjects were divergent. Subject n. 17, characterized by severe conditions, was SARS-CoV-2 positive in different specimens, including maternal plasma, vagina as well as umbilical cord plasma and placenta. In this case, we hypothesize that the virus spread around the body through the bloodstream, reaching the vagina and the placenta, finally infecting the fetus. Indeed, the nasopharyngeal swab of her newborn collected upon delivery resulted positive. Notably, subject n. 17 was the only one to deliver prematurely at week 34. Prematurity was indeed reported to be more frequent in SARS-CoV-2 infected patients^8^. We hypothesize that this might be related to the inflammatory status, as a consequence of the viral infection; alternatively, this could have been the result of a pre-existing condition that triggered the premature delivery and facilitated viral entry through the placenta. SARS-CoV-2 positivity of umbilical cord plasma from subject’s 17 newborn proves an *in-utero* transmission. In the same woman, vagina was found positive for SARS-CoV-2. Since the presence of the virus in cord blood indicates an *in-utero*-transmission prior to delivery, we cannot speculate about the risk of acquiring the virus during vaginal delivery in this case. However, we cannot exclude the possibility of viral intrapartum infection when the virus is present in the vagina. Subject n. 17 delivered 6 days after the first COVID-19 diagnosis. Probably due to the short span of ΔT1-T0 time, specific anti-SARS-CoV-2 IgM were not detected in umbilical cord blood. Conversely, subject n. 25 who manifested mild symptoms was SARS-CoV-2 negative in all the biological samples analyzed (maternal plasma, vagina, umbilical cord plasma), but the placenta. However, her newborn had a SARS-CoV-2 positive nasopharyngeal swab at birth and both SARS-CoV-2 specific IgM and IgG were detected in umbilical cord plasma. Although still controversial^25^, the presence of anti-SARS-CoV-2 IgM strongly suggests SARS-CoV-2 infection *in-utero*. Of note, the positivity of the newborns’ nasopharyngeal swab was not sustained over time, as the following tests were negative. We detected IgM and IgG in maternal plasma as well. This is consistent with the span of time occurred between COVID-19 diagnosis and delivery (17 days), where the median of detection of specific IgM/IgG is 13 days^26^.

We analyzed the milk collected at T2 as well. We detected the presence of SARS-CoV-2 RNA in one case only (subject n.1), who was a severe case. This is consistent with what previously reported^27^. However, further studies are required to assess whether this represents an infectious and replicative virus or not. Although precautions were adopted, we cannot exclude a contamination of the sample by other maternal positive sites. Moreover, we tested milk specimens for the presence of specific anti-SARS-CoV-2 IgM and IgG. We were able to detect IgM in subject n.1 only. It was previously reported that the absence of IgM and IgG in the milk is not uncommon, especially in the case of respiratory viruses^28^. A recent study showed the high sensitivity and specificity of iFlash automated system for antibodies detection^23^. However, this methodology has been adapted for detection of antibodies in milk and the sensitivity may be attenuated on this particular specimen.

Further studies are needed to ascertain long-term outcomes and potential intrauterine vertical transmission in pregnant women infected in the first or second trimester. This observation is even more relevant considering that the temporal and spatial expression of the main SARS-CoV-2 receptor, ACE2, has been reported to change significantly in maternal-fetal interface tissues in the different trimesters^21,29^. We can speculate on the possibility that ACE2 modulation could be directly linked to placenta susceptibility to SARS-CoV-2 infection. Alternatively, we can reason on the possibility that due to altered permeability/damages of the placenta, probably secondary to an inflammatory status, SARS-CoV-2 is able to bypass the placental barrier and reach foetal blood. This issue still remains to be addressed and further investigated.

As several lines of evidence indicate that systemic maternal infection and consequent inflammation contribute to disruption of placenta development/function and possibly favour viral vertical transmission ^30,31^, we decided to profile the inflammatory status of four selected patients at both local (placenta) and systemic (maternal and fetal) level. Results obtained by different molecular approaches (RNA expression and protein secretion) give us the same take-home message by showing a trend of generalized immune activation in those patients (17 and 25), who were SARS-CoV-2 positive at delivery and, according to the viral-immunological analyses, infected their neonates *in utero*. Unexpectedly, this hyper-activation status was far more evident in SARS-CoV-2–negative biological samples (placenta biopsy, maternal and umbilical cord plasma) from subject n. 25, compared to subject n. 17, who displayed severe clinical condition. A plausible explanation to this apparent inconsistency stems from the observation that subjects n. 17 was undergoing a cortisone prophylaxis during the antepartum period that could have weakened the synthesis and release of inflammatory cytokines/chemokines. Among the inflammatory factors, whose expression was evidently increased in in both placenta and cord blood samples from subjects n. 17 and 25, the chemokines, CXCL10, CXCL8, CCL5, CCL3, CCL2, could have played a major role in favouring vertical transmission. Indeed, they could have created a chemotactic gradient between villi and the inter-villous space, where maternal lymphocyte circulate, thus favouring viral dissemination^32^. To perform such molecular analyses, only four subjects (CTR-, n. 17, n. 25 and n. 31) were chosen due to their peculiarities. However, further experiments are envisaged, in order to confirm this distinctive profile.

In conclusion, for the first time SARS-CoV-2 was detected in umbilical cord plasma, strongly suggesting that *in-utero* mother-to-child transmission is possible and apparently related to a high maternal inflammatory state. Although further studies are needed, this should be taken into consideration in the management of COVID-19 pregnant women.

## Data Availability

All data will be available upon request

## Acknowledgements

We would like to thank Paolo Quaini, Francesco Leone, Federica Fusè, Irma Saulle, Claudia Fusetti, Margherita Longo, Alberto Rizzo, Francesca Romeri, Federica Brunetti and Francesca Sabbatini for their support and contribution to the project.

## Authors’ contributions

VS conceived the presented idea. MB further developed the project with the help of CF. VS, IC, PV, AS, FP, CC and SC performed subject enrolment and clinical management, as well as samples collection. IB was involved in sample collection and management. CF and MB conceived, planned, performed and analysed the experiments on SARS-CoV-2 detection on plasma, biopsies and vaginal swab, and experiments on the inflammatory response. DT helped with the interpretation of the data. DM and AM conceived, planned and performed the experiments on specific antibody detection and experiments on milk, under the supervision of MG. CF, MB and VS discussed the data and wrote the manuscript. MC, IC and PV critically reviewed the manuscript. CF and MB equally contributed to the manuscript.

